# Functional Activity of TDP 43: A Direct Biomarker for ALS

**DOI:** 10.64898/2026.05.04.26352054

**Authors:** Kirti Shila Sonkar, Vito Levi D’Ancona, Jade Cramp, Hannah Shilling, Ellie Giles, Tyler Howell-Bray, Becky Fillingham, Merit E. Cudkowicz, Avindra Nath, Jeffrey D. Rothstein, Robert Bowser, Barbara Borroni, Alessandro Padovani, James D. Berry, Ghazaleh Sadri Vakili, Emanuele Buratti, Ian P. Thrippleton

## Abstract

TDP-43 dysfunction is a defining feature of amyotrophic lateral sclerosis (ALS), yet no biofluid biomarker directly measures its functional activity. We developed a serum-based homogeneous time-resolved FRET (hTR-FRET) assay that quantifies TDP-43 RNA binding activity using synthetic UU-rich RNA probes. We analyzed 1,080 serum samples from controls, sporadic ALS, and genetic subgroups (C9orf72, SOD1) across multiple biorepositories. Cross-sectionally, TDP-43 functional activity was elevated in ALS (mean 390 a.u.) versus controls (304 a.u.), yielding AUC = 0.79. Genotype means were 392 a.u. (sporadic), 382 a.u. (C9orf72), and 323 a.u. (SOD1); a 366 a.u. threshold achieved 95% specificity against controls. Longitudinally, Target ALS showed a modest but significant inverse correlation between TDP-43 activity and ALSFRS-R, while other cohorts exhibited similar non-significant trends. Elevated signal in serum likely reflects increased extracellular, probe-competent TDP-43 species. This assay provides a proof-of-concept platform for the direct functional measurement of probe-competent TDP-43 species in serum. While it demonstrates moderate group-level discrimination, individual diagnostic performance requires prospective validation. The assay may support exploratory applications in genotype stratification and progression monitoring in future clinical studies.

## 1. Introduction

TAR DNA-binding protein 43 (TDP-43) is a ubiquitously expressed, multifunctional DNA/RNA binding protein critical for essential cellular processes, including transcriptional regulation, pre-mRNA splicing, mRNA stability, and microRNA biogenesis [1-5]. Its function is critically dependent on its distinct domain structure, featuring two RNA binding domains (RBD) that mediate high-affinity binding to UU-rich RNA sequences, and a C-terminal domain that is intrinsically disordered and prone to aggregation [2,6]. Under physiological conditions, TDP-43 is predominantly nuclear but shuttles between the nucleus and cytoplasm to perform its duties in RNA metabolism [7,8]. The central role of TDP-43 in neurodegeneration was unveiled when it was identified as the primary constituent of the pathological cytoplasmic inclusions in the majority of Amyotrophic Lateral Sclerosis (ALS) cases and a significant subset of Frontotemporal Dementia (FTD) cases [9] and recently reviewed by [3,10]. This pathology, termed TDP-43 proteinopathy, is characterized by the nuclear depletion of TDP-43, its cytoplasmic mislocalization, hyper-phosphorylation, and cleavage into insoluble aggregates [7]. These aggregates are not merely a hallmark of disease but are directly implicated in pathogenic cascades through a loss of normal nuclear function and a potential gain of toxic function in the cytoplasm [1]. The critical loss of TDP-43’s splicing repression function, for instance, has been shown to lead to aberrant cryptic exon inclusion in transcripts like UNC13A and STMN2, genes vital for neuronal survival [11],[7]. TDP-43 pathology is now recognized as a common feature across a spectrum of disorders, like ALS, FTD, Alzheimer’s Disease and Limbic-predominant Age-related TDP-43 Encephalopathy [4,8,10]. Despite its clear pathological significance, a significant gap persists in our ability to measure the functional state of TDP-43 in vivo during a patient’s life [12]. While emerging biomarkers, such as cryptic peptides (e.g., HDGFL2), are beginning to reflect TDP-43 dysfunction, they are indirect readouts and not a direct measure of the protein’s core RNA-binding function.

While conventional immunoassays and modern high-throughput proteomic platforms excel at quantifying static protein abundance or specific post-translational modifications, [1,8] they are inherently blind to biochemical competence. In the case of TDP-43, pathogenicity is driven not merely by its presence, but by its loss of nuclear RNA-binding function [1]. and cytoplasmic mislocalization. Therefore, a biomarker that directly captures probe-competent TDP-43 activity addresses a critical gap in measuring the protein’s functional state in vivo. Moreover, the use of different biofluid matrices, particularly plasma versus serum, has led to inconsistent results, with evidence suggesting that plasma anticoagulants can interfere with TDP-43 measurements [13]. A biomarker that detects extracellular, probe-competent TDP-43 species in serum would therefore address a critical gap. The assay described here measures TDP-43’s ability to recognize and bind synthetic UU-rich RNA motifs in a biofluid context; it does not purport to reflect nuclear RNA-binding function or restored splicing competence.

Foundational studies defined the UU-rich sequence grammar that underlies TDP-43’s RNA specificity, providing a molecular blueprint for a function-based assay [14]. Building on this knowledge, we constructed a nucleotide sequence fulfilling the TDP-43 motif requirements. We established a homogeneous hTR-FRET assay in which the TDP-43 motif dependent probe generates a measurable signal that reflects its binding activity in context (Fig1a). By introducing a direct functional biomarker for TDP-43, the work described here aims to bridge the gap between molecular pathology and clinical application, supporting applications in population-level discrimination of ALS from controls and exploratory longitudinal trends. The ALS Functional Rating Scale Revised (ALSFRS R) is a validated clinical tool used to quantify motor function decline and disease progression, which we utilized to correlate biochemical changes with clinical severity.

**Figure 1.**
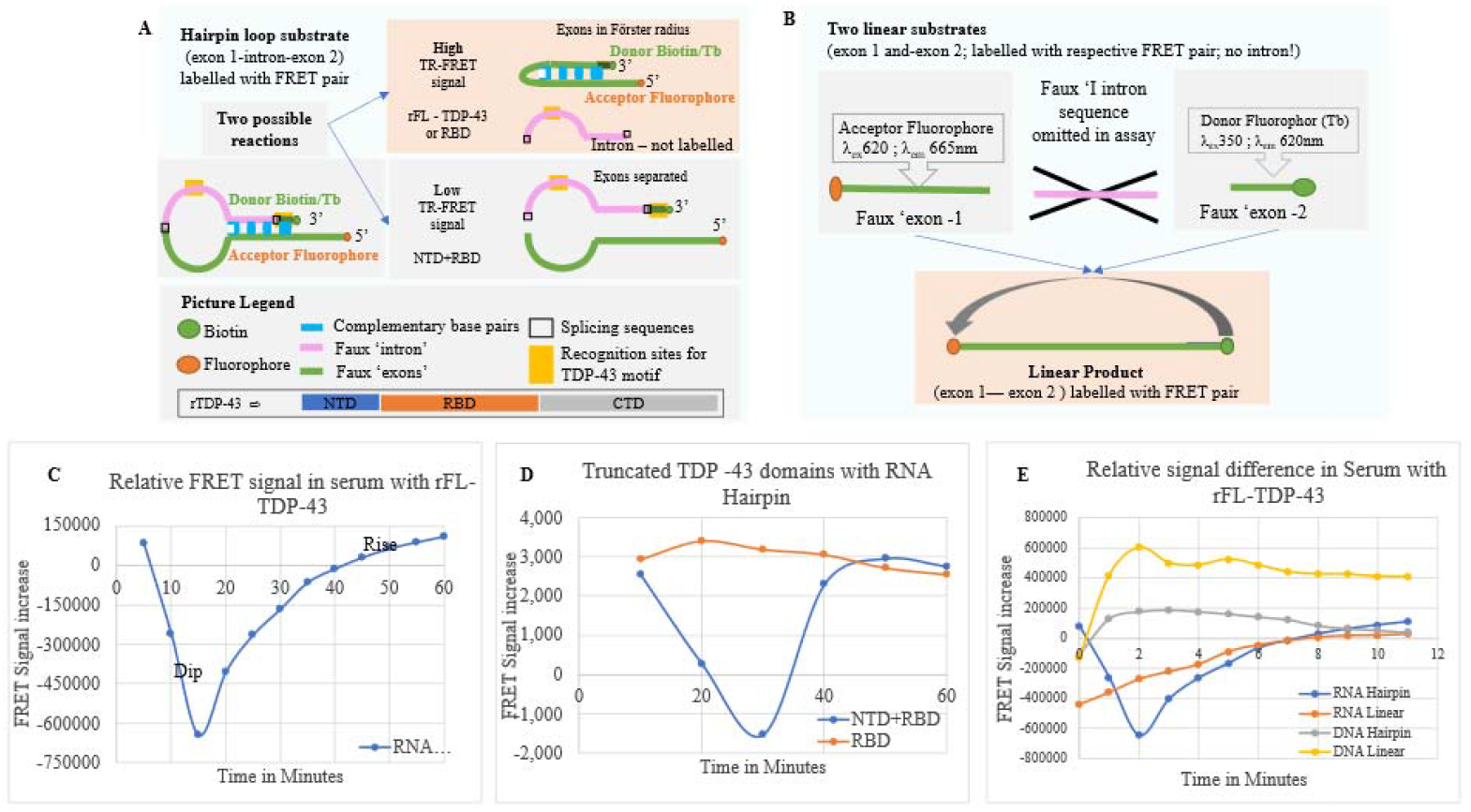
Validation and characterization of a hTR-FRET Assay for TDP-43 functional activity. (A) Schematic of the hairpin-loop substrate assay. The substrate comprises exon 1, a faux intron, and exon 2, labeled with a FRET pair. Intact hairpins maintain donor and acceptor fluorophores within the Förster radius, generating a high baseline TR-FRET signal. Cleavage by TDP-43 physically separates the exons, distancing the fluorophores and quenching the signal. The diagram also illustrates the domain architecture of recombinant TDP-43 (NTD, RBD, CTD) and the two possible reaction outcomes. (B) Linear bridging assay design. To isolate probe-bridging activity, the assay was simplified to two independent synthetic probes: a “faux exon 1” (acceptor fluorophore) and a “faux exon 2” (donor fluorophore). The intron sequence is omitted to enhance substrate stability. In solution, these free substrates cannot generate a FRET signal. A signal is produced exclusively when functional TDP-43 bridges the two probes, bringing the fluorophores within the Förster radius. (C) Kinetic profile of the hairpin assay with recombinant full-length TDP-43 (rFL-TDP-43). The assay exhibits a characteristic biphasic “dip-then-rise” signature over a 60-minute window. The initial dip reflects transient FRET pair separation, while the subsequent monotonic increase indicates donor-acceptor re-association driven by TDP-43-mediated probe alignment. (D) Domain mapping of TDP-43 activity using RNA hairpin substrates. Constructs containing the N-terminal domain (NTD) fused to the RNA-binding domain (RBD) retain the full biphasic signature. In contrast, the RBD domain alone supports only the signal rise phase, indicating that the NTD is required for the initial structural rearrangement or dip phase. (E) Substrate specificity analysis comparing RNA versus DNA probes. Only RNA substrates containing canonical TDP-43 binding motifs elicit the characteristic dip-then-rise kinetic profile. DNA probes fail to produce the rise phase, confirming that signal generation is strictly RNA-dependent and requires full-length TDP-43. Data represent mean ± SEM of technical triplicates. Time (minutes) is shown on the x axis; normalized hTR FRET ratio (665/620 nm, a.u.) on the y axis.

## 2. Materials and Methods

### 2.1 Study Design

Analytical decisions were pre-specified prior to unblinding: single-hypothesis testing (TDP-43 RNA-bridging competence as the outcome), run triplicate protocol with CV >15% exclusion, and pre-defined LoB/LoD thresholds (see Section 2.6). All experiments prospectively tested a single hypothesis: that a synthetic UU rich RNA substrate could convert the functional state of TDP 43 in human serum into a quantifiable optical signal.

### 2.2 Serum samples

The study integrates data from multiple sources as mentioned in table 1. ALS samples from NEALS (AIM, Bio1, Bio2, Bio3, ALS biorepository), NIH/NINDS repositories, the Spedali Civili Hospital of Brescia clinical biobank, ANSWER ALS, Target ALS, AD and FTD samples from iSpecimen totaling 1080 serum samples. Each sample was processed for the hTR-FRET analysis under uniform analytical conditions, and every plate incorporated reference controls to ensure cross-site comparability. The assay design is mechanism-driven, leveraging known TDP-43 RNA-binding domain geometry and FRET physics to convert functional RNA engagement into a quantifiable optical signal.

**Table 1.** Serum Distribution by Diagnostic Category: Tabulated breakdown of diagnostic categories, subgroup genotypes, sample counts, sources, and key notes on cohort composition.

| Diagnostic Category | Specific Group / Genotype | Sample Count (Total N=1080) | Source(s) |
| --- | --- | --- | --- |
| Control | Healthy Controls | 145 | Research Donors from established biobanks (NEALS, NIH/NINDS, iSpecimen) |
| Amyotrophic Lateral Sclerosis (ALS) | Sporadic ALS (sALS) | 839 | NEALS (AIM, Bio1, Bio2, Bio3, ALS biorepository), NIH/NINDS, Spedali Civili Hospital of Brescia, ANSWER ALS, TARGET ALS. |
|  | Familial ALS (fALS):<br>• C9ORF72-ALS (n=51)<br>• SOD1-ALS (n=15) | 66 |  |
| Other Neurodegenerative Diseases (Comparator Group) | Frontotemporal Dementia (FTD) and FTD-GRN | 10 | iSpecimen |
|  | Alzheimer’s Disease (AD) | 20 |  |
| Note: Detailed demographic characteristics (age, sex, ethnicity, race) and longitudinal metrics for the ANSWER ALS cohort are provided in Table S1. |  |  |  |

### 2.3 IRB and sample collection information

Samples included in this analysis were drawn from prospectively designed biorepositories and longitudinal studies, but analyzed retrospectively for this cross-sectional/exploratory study, spanning a mixed population and different cohort across different geographies. All sample collections were carried out under protocols approved by the MassGeneral Brigham Institutional Review Board (IRB), Barrow Neurological Institute IRB, NIH/NINDS and Spedali Civili Hospital of Brescia, Italy, relevant local IRBs at each relevant participating study site, and TARGET ALS (samples from 2 patients were considered compromised and not included in our analysis). Prior to undergoing any data collection or study procedures, all participants underwent an informed consent process and provided written informed consent.

#### 2.3.1 Human plasma and serum samples

A random sampling of de-identified plasma and serum from sALS (102 sALS) and healthy controls (10 HC) were obtained from the Northeast Amyotrophic Lateral Sclerosis Consortium (NEALS) Biofluid Repository at Mass General Brigham and the Barrow Neurological Institute, a shared repository containing more than 100,000 biofluid samples from people with ALS and healthy controls for use by the ALS research community. The samples and associated clinical data were collected between 2008 and 2023 from participants in multi-site research studies. Of the 102 sALS, 94 had plasma and serum samples available from two to three visits; one had serum only and two had plasma only available. Serum was also provided from a single visit for healthy controls. Across all studies, serum samples were collected using standard red-top tubes (no SST gel additives), allowed to clot fully at room temperature for 30 minutes, and then centrifuged (1,750 × g, 10 min). Supernatant was carefully aliquoted without disturbing the platelet/cell pellet. The 10 plasma samples were collected using EDTA tubes and blood for serum using standard red top tubes and centrifuged immediately. Vacutainers were centrifuged at 1750 g for 10 min, supernatants aliquoted into cryovials and frozen at -70°C to -80°C within two hours of collection. Samples were stored below -70°C, with no freeze-thaw cycles. Residual platelet-derived TDP-43 contamination cannot be excluded and is acknowledged as a limitation. Future studies will evaluate SST vs. red-top tube impacts on assay signal.

#### 2.3.2 Serum vs Plasma

Matrix Selection: Serum was selected over plasma based on mechanistic considerations. Plasma contains polymeric fibrinogen and anticoagulants (EDTA, heparin, citrate) additives that bind TDP-43 and attenuate free protein **pools [**15,23] and introduce non-specific FRET quenching and ionic variability that disrupt donor–acceptor coupling. Serum, by contrast, better represents the physiologically relevant soluble fraction of TDP-43 released from cells and exosomes. Consistent with NEALS and ANSWER ALS protocols, all samples in this study were serum derived to avoid matrix related artifacts and maintain compatibility with reference datasets. Empirical testing of matched serum–plasma pairs from ten donors confirmed this choice: serum produced stable biphasic kinetic traces, whereas plasma yielded flat or erratic profiles (data not shown). Taken together, serum was adopted as the exclusive matrix.

#### 2.3.3 Specificity Validation

Specificity was validated through orthogonal experimental axes: (1) chemistry dependence (RNA but not DNA substrates produced the biphasic kinetic trace), and (2) protein identity (full-length (1-414; TDP-43 FL) and NTD+RBD construct showed dip and rise activity, whereas RBD-only construct (without N terminus) showed only the rise phase. With this pattern, we can conclude that TDP-43 N-terminus domain is responsible for the initial dip phase. This validation established that the fluorescence increase cannot arise from random molecular crowding or nonspecific nucleoprotein complexes.

### 2.4 RNA Substrate Design

#### 2.4.1 Rationale

TDP-43 binds RNA through its tandem RNA binding domain (RBD) that sandwiches UU-rich sequences into a shallow hydrophobic cleft [20]. These insights were used to design a pair of synthetic RNA exons, in which one carries a UU motif, flanked by stabilizing spacers that mimic natural exon junctions. Both oligonucleotides were 2′-O-methyl modified to improve nuclease resistance without altering hydrogen-bonding geometry.

#### 2.4.2 Architecture and Labeling

Probe A carries a red-emitting acceptor fluorophore (excitation ≈ 620 nm; emission ≈ 665 nm); Probe B is biotinylated and couples via streptavidin to a terbium cryptate donor (excitation ≈ 350 nm; emission ≈ 620 nm). When TDP-43 bridges the UU motifs, the probes move within the Förster radius (R_0 ≈ 7 nm), enabling distance-dependent energy **transfer [**24]. Using the Förster relationship (E = 1 /[1 + (r /R0)6], this setup provides a homogeneous, physics-driven assay suitable for detecting analytes in impure biological samples.

#### 2.4.3 Control Substrates

In addition to the UU-based probe, three categories of controls were employed: DNA versions of the probes (lacking 2′-OH) to test chemistry dependence and scrambled competitors (“cold” oligos) to test sequence dependence and truncated proteins.

### 2.5 hTR-FRET Detection Physics

Time-resolved Förster resonance energy transfer (hTR-FRET) was selected for its ability to separate long-lived donor emission from short-lived biofluid autofluorescence, enabling high-sensitivity serum assays in the pg/mL–fg/mL range.

#### 2.5.1 Physical Basis

FRET transfers energy from an excited donor to an acceptor via dipole–dipole interaction, with efficiency highly dependent on proximity [24]. To achieve sensitive detection in complex fluids, we employed lanthanide-based probes in a homogeneous format. A Streptavidin–Terbium cryptate donor was chosen for its millisecond-scale life-time (≈1.5–1.6 ms), compared to nanosecond lifetimes of endogenous fluorophores. By delaying detection (Δt = 60 µs) and integrating over a 400 µs gate, background fluorescence was effectively eliminated. Signal was quantified as the ratio of acceptor to donor emission (665 nm /620 nm), normalized to the per-plate healthy pooled serum (HPS) control mean, and reported in arbitrary units (a.u.). Positive control: 50 nM recombinant FL-TDP-43 (expected 420–480 a.u.); negative control: probe blank without protein (expected <200 a.u.). These benchmarks were included on every plate to define assay dynamic range and normalization reference. Fluorescence lifetime acquisition is not available on the BMG VANTAstar platform used. While lifetime-based FRET analysis would offer higher precision, intensity-based ratiometric detection was employed due to platform constraints. Error propagation for the FRET ratio follows standard ratiometric analysis: ΔR/R = √[(ΔI_665_/I_665_) ^2^ + (ΔI_620_/I_620_) ^2^], which forms the basis of the reported intra-assay CV.

#### 2.5.2 Instrumentation

All assays were performed on BMG Labtech ClARIOstar Plus and VANTAstar F readers in TR/F mode with the following parameters: excitation at 350 ± 10 nm, emission 1. at 620 ± 10 nm, and emission 2 at 665± 10 nm with delay of 60 µs, integration at 400 µs, 20 flashes per read, 25 °C, top optics. Plates were Corning 384-well low-volume white microplates, with each sample run in triplicate and column-wise inclusion of healthy pooled serum (HPS) and blank controls.

### 2.6 Assay Workflow and Quality Control

Each sample run consisted of: 5 µL triplicate wells per sample (15 µL total); Two incubation phases at 25 °C for 1□hour each to allow equilibrium binding and conformational closure. Triplicates with a coefficient of variation (CV) > 15% were automatically excluded. Analytical thresholds were calculated as: limit of blank (LoB) = mean blank + 1.645 × SD blank; limit of detection (LoD) = LoB + 1.645 × SD low. Across 28 plates, intra-assay CV averaged 4.4% and inter-plate CV 15%, meeting or exceeding clinical laboratory precision standards (Figure S1). Plates failing control criteria were removed from analysis.

### 2.7 Statistical Analyses

Data sets were processed in R 4.3 using packages tidyverse, rstatix, and FactoMineR. Primary outcome: mean hTR-FRET values (a.u.) per sample. Group comparisons employed Welch’s t-tests; effect sizes expressed as Cohen’s d. Precision: CV = 100 × SD /mean.

### 2.8 Longitudinal Cohort Designs

To assess temporal stability and progression, we analysed serial serum samples from NEALS, Target ALS, and ANSWER ALS repositories. Participants (ALS) had 2–6 visits over 6–24 months, linked by unique identifiers. All samples were processed using the same hTR-FRET protocol and calibration curves. Survival data were derived from biorepository clinical databases and linked via unique identifiers. Incomplete follow-up is acknowledged as a limitation. Changes in TDP-43 activity were expressed as ΔFRET = FRET (visit n) – FRET (baseline, visit 1). Sensitivity analyses using visit-to-visit changes (FRET (visit n) – FRET (visit n−1)) yielded similar conclusions. Statistical analysis used paired t-tests for within-subject comparisons and mixed-effects linear models to estimate progression slopes by diagnostic group. All assay runs were performed blinded to diagnostic status. Cases and controls were randomly distributed across plates to minimize batch effects.

## 3. Results

### 3.1. A novel hTR-FRET assay measures the functional activity of TDP-43

To quantify the functional RNA-binding state of TDP-43 in biofluids, we developed a serum-based, homogeneous time-resolved Förster resonance energy transfer (hTR-FRET) assay. The primary biomarker of interest is the amplitude of the DELTA-FRET signal (DELTA-FRET = FRET (visit) - FRET (baseline)), which reflects probe-competent TDP-43 bridging activity. For assay designing initially, we utilized a hairpin-loop substrate fulfilling TDP-43 motif requirements, schematically represented in Fig. 1a. This substrate consists of exon 1, a faux intron, and exon 2, labeled with a FRET pair. In this configuration, the hairpin structure maintains the donor (Biotin/Tb) and acceptor fluorophores within the Förster radius, generating a high baseline TR-FRET signal. Cleavage of the hairpin loop physically separates the FRET pair, liberating the exons into solution where FRET energy transfer is physically impossible, thereby reducing the detected signal. However, the hairpin substrate produced a biphasic “dip-then-rise” kinetic signature (Fig. 1c). The initial dip corresponds to a transient signal decrease due to FRET pair separation, while the subsequent monotonic increase reflects the re-association of FRET partners within the Förster radius. This rise phase was observed only in the presence of full-length TDP-43. To ensure unambiguous detection of this bridging activity and minimize potential serum nuclease interference, we simplified the substrate architecture from the Hairpin loop substrate to a linear substrate architecture (Fig. 1b).

This final design employs two synthetic probes: a “faux exon 1” labeled with the acceptor fluorophore (λex 620 nm; λem 665 nm) and a “faux exon 2” labeled with the donor (Terbium; λex 350 nm; λem 620 nm). Crucially, the faux intron sequence is omitted to enhance stability. In this linear setup, free substrates in solution cannot generate a FRET signal. A signal is generated exclusively when functional TDP-43 bridges these two probes, bringing the donor and acceptor within the Förster radius (r→R0 ≈ 7 nm). This proximity-dependent phase reflects donor–acceptor coupling mediated by probe alignment, serving as a functional fingerprint of active TDP-43–RNA engagement.

For kinetic characterization and domain mapping we did the second phase of the incubation and plateauing by 60 min emerges only in the presence of probe-competent TDP-43. Kinetic modeling confirmed this represents a first-order proximity process, with the amplitude of ΔFRET correlating directly with TDP-43 functional levels in serum. No non-TDP-43 proteins fit the observed curve. To elucidate the molecular requirements for this activity, we performed domain mapping (Fig. 1d). Constructs containing the N-terminal domain (NTD) together with the RNA-binding domain (RBD) retained both the dip and rise activities. Intriguingly, the RBD domain alone supported only the signal rise activity and failed to reproduce the full biphasic signature. This highlights a potential role of the NTD sequence in the liquid-liquid phase separation (LLPS) or structural rearrangements of the mix. Specificity Validation Additional validation confirmed the assay’s RNA specificity. In the presence of full-length TDP-43, RNA substrates containing canonical UU motifs produced the characteristic dip-then-rise signature, whereas DNA probes failed to elicit the rise phase (Fig. 1e). While RNA hairpins generated the distinct dip followed by a gradual increase consistent with secondary structure stabilization, DNA hairpins showed only linear binding. These results confirm that the observed hTR-FRET value is RNA-specific and dependent on full-length TDP-43.

### 3.2 TDP-43 hTR-FRET functional activity as a diagnostic biomarker for ALS

We applied the hTR-FRET assay to a large cross-sectional cohort of serum samples. TDP-43 functional activity was significantly elevated in ALS patients (n = 905; mean = 390 ± 99 a.u.) compared to healthy controls (n = 145; mean = 304 ± 36 a.u.; p < 0.0001, Welch’s t-test) (Table 2).

**Table 2.** Summary of cross-sectional TDP-43 hTR-FRET values.

|  | Number | Mean TDP-43* | vs Healthy | vs ALS |
| --- | --- | --- | --- | --- |
| Healthy Control | 145 | $304 \pm 36$ | | |
| ALS (sALS) | 839 | $392 \pm 99$ | $p < 0.001$ *** | |
| C9orf72-ALS | 51 | $382 \pm 87$ | $p < 0.001$ *** | |
| SOD1-ALS | 15 | $323 \pm 105$ | $p < 0.001$ *** | |
| FTD | 10 | $458 \pm 159$ | $p < 0.001$ *** | $p = 0.086$ (ns) |
| AD Fast | 10 | $443 \pm 74$ | $p < 0.001$ *** | $p = 0.059$ (trend) |
| AD Slow | 10 | $478 \pm 200$ | $p < 0.001$ *** | $p = 0.256$ (ns) |

\* Values reported as Mean ± SD.

This difference demonstrated with a large effect size (Cohen’s d = 0.93) and good classification accuracy (AUC = 0.79, 95% CI: 0.76–0.82; p < 0.001). TDP-43 functional activity was quantified using the hTR-FRET assay, which demonstrated intra-assay precision of 4.4% CV and inter-plate CV of 15% (see Section 2.6). The ROC curve is presented in Figure S1. Baseline hTR-FRET values differed significantly across diagnostic groups (p < 0.0001). Cross sectionally, TDP 43 activity discriminated ALS from healthy controls with a large effect size. It is important to note that population-level statistical separation (AUC, effect size) differs from individual diagnostic performance. Predictive values (PPV/NPV) require prospective study designs with known disease prevalence and cannot be derived from this retrospective cohort. The observed population-level separation warrants further validation before clinical application.

### 3.3 Genotype-Specific TDP-43 functional activity: Genetically Confirmed vs. Sporadic ALS

Table 2 summarizes hTR-FRET values across diagnostic groups. Healthy controls (n = 145) exhibited a mean of 304 ± 36 a.u., while sALS cases showed significantly higher activity (392 ± 99 a.u.; p < 0.001). Genetically confirmed ALS subtypes demonstrated distinct profiles: C9orf72 ALS averaged 382 ± 87 a.u., and SOD1 ALS showed lower activity (323 ± 105 a.u.), all pre tofersen treatment. These findings suggest genotype-specific variation in extracellular, probe competent TDP 43 species detectable by the assay. Comparator groups exhibited values that overlapped with or exceeded ALS: FTD (458 ± 159 a.u.), AD-fast (443 ± 74 a.u.), and AD-slow (478 ± 200 a.u.), though these cohorts were small (n = 10–20). While differences versus ALS were not statistically significant (p = 0.059–0.256), the trend indicates potential disease-specific patterns requiring validation in larger cohorts.

### 3.4 Diagnostic Thresholds and Distribution Analysis of TDP-43 hTR-FRET Values

Figure 2 illustrates the distribution of hTR-FRET values in ALS patients (n = 905) versus healthy controls (n = 145). A cutoff of 366 a.u. achieves 95% specificity, effectively separating ALS from controls. Kernel density curves highlight the distinct profiles: controls exhibit a narrow, approximately normal distribution centered at lower values, while ALS shows a broader, right-shifted distribution with higher means, reflecting elevated TDP-43 functional activity. Control-derived thresholds define specificity against healthy controls for research purposes: 366 a.u. (95% specificity), 338 a.u. (90% specificity), 311 a.u. (70% specificity), and median 303 a.u. Sensitivity at each threshold is reported in Table 2. Clinical application requires prospective validation. The large effect size (Cohen’s d = 0.93) and group separation underscore the assay’s potential for population-level discrimination in research settings. Cutoff selection should be guided by the intended application (e.g., screening vs. confirmatory) in future prospective studies.

**Figure 2.**
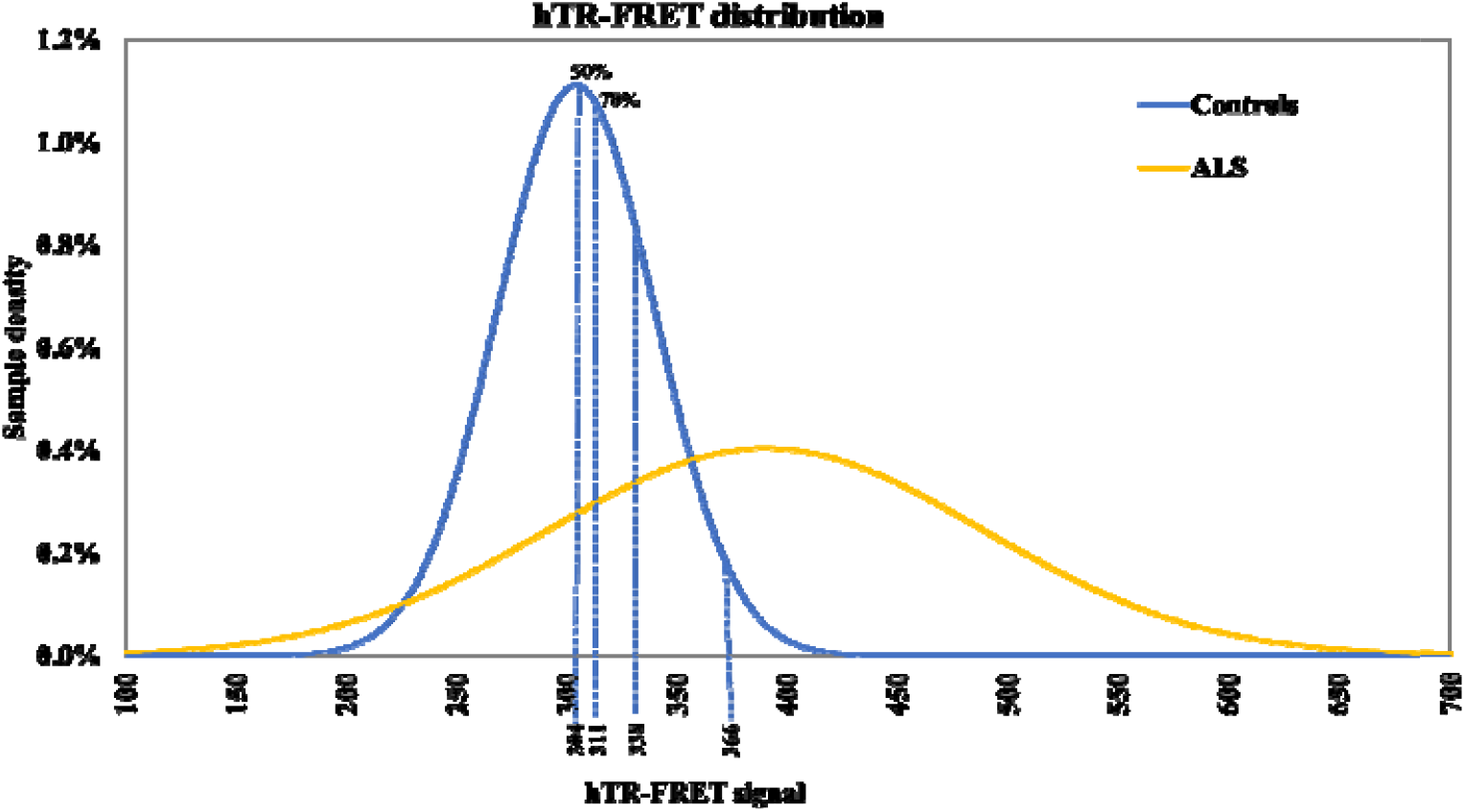
Distribution and diagnostic performance of TDP-43 hTR-FRET values in ALS and controls: ALS distribution shows the distribution of TDP-43 hTR-FRET value in ALS patients (n=905) compared to healthy controls (n=145). A 366 a.u. TDP-43 FRET value serves as the cutoff for 95% specificity. The large effect size (Cohen’s d=0.93) indicates good separation between disease and healthy states, supporting the potential for population-level discrimination in research settings.

### 3.5 Diagnostic Group Comparison of TDP-43 hTR-FRET Activity

Cross-sectional analysis of hTR-FRET TDP-43 signal revealed statistically significant group-level differences across diagnostic categories, with significant overlap between individual distributions (Figure 3). Healthy controls clustered tightly at lower values, establishing a baseline of minimal TDP-43 function activity. In contrast, ALS samples were significantly right-shifted and more dispersed, indicating elevated and variable activity. The 95% control separation threshold (366 a.u.) effectively distinguished ALS from hibit lower activity than sporadic ALS, consistent with potential genotype-specific differences in RNA dysregulation. AD and FTD samples showed higher values, supporting the assay’s ability to detect TDP-43-related dysfunction across neurodegenerative conditions.

**Figure 3.**
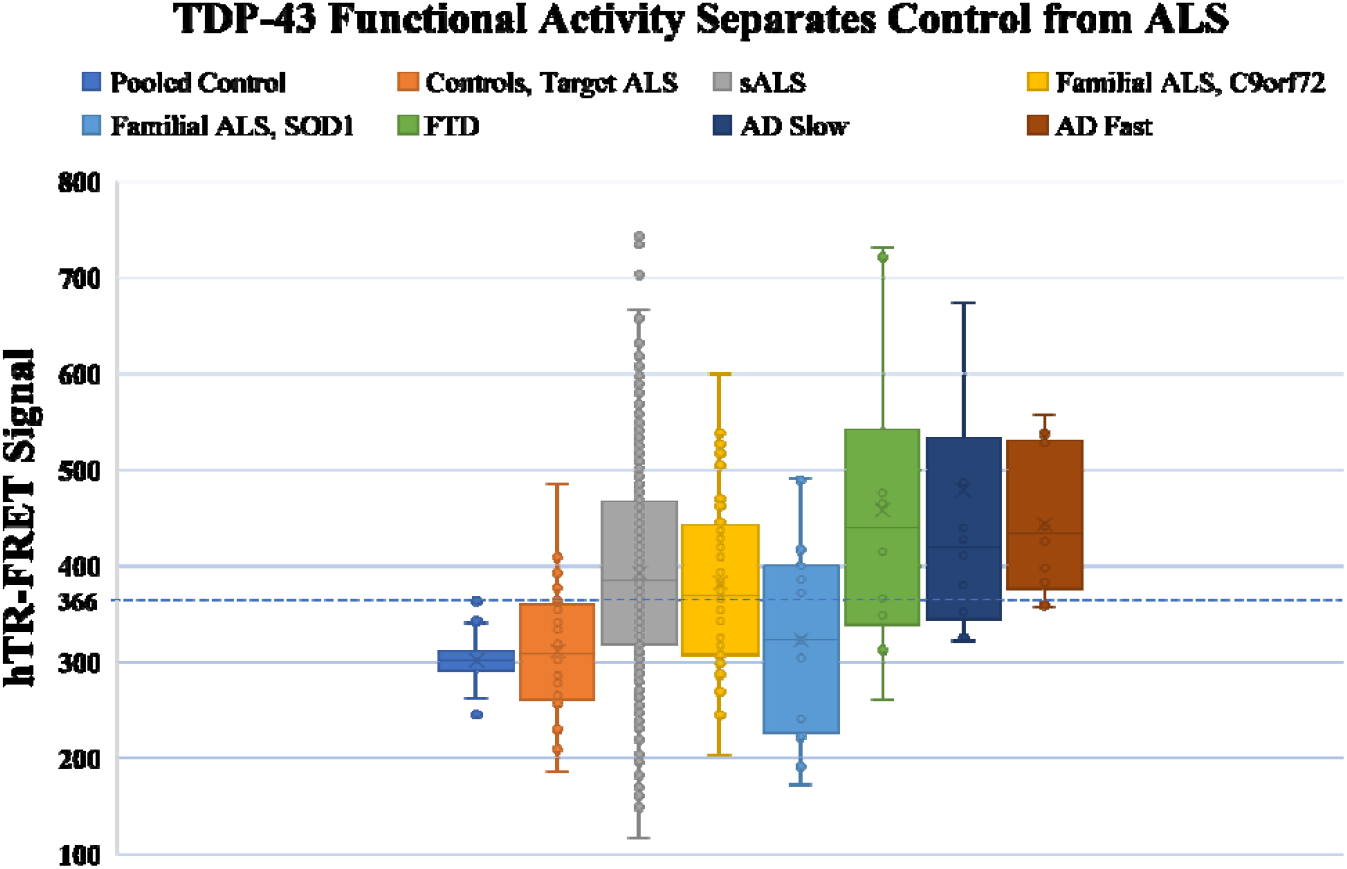
Cross-sectional hTR-FRET TDP-43 functional activity across Diagnostic Group: Boxplots and overlaid kernel density estimate for Healthy Controls, Probable ALS, Definite ALS, and Other neurodegenerative diseases (AD/FTD). Each box shows median and interquartile range; whiskers extend to 1.5× IQR and individual points represent samples that passed triplicate QC. Annotation: dashed vertical line at 366 a.u. marks the 95% control separation threshold.

### 3.6 Longitudinal Analysis Shows High Variability and General Stability

Longitudinal analyses of serial serum samples revealed variable slopes of TDP-43 hTR-FRET functional activity across cohorts. Across all cohorts, mean changes per visit were modest and did not reach statistical significance, reflecting substantial inter-individual variability. In smaller cohorts such as NEALS Bio01 (9 subjects, 22 measurements; slope −1.16 ± 31.77 a.u./visit, p = 0.920), NEALS Bio03 (17 subjects, 48 measurements; slope −13.56 ± 124.99, p = 0.670), and NEALS AIM (25 subjects, 64 measurements; slope −6.35 ± 47.13, p = 0.516), slopes were near zero or negative. By contrast, larger cohorts demonstrated positive mean slopes suggestive of gradual increases in TDP-43 function activity over time: NEALS Bio02 (36 subjects, 103 measurements; slope 9.59 ± 38.46, p = 0.149), Answer ALS (112 subjects, 341 measurements; slope 7.87 ± 54.13, p = 0.128), and Target ALS (18 subjects, 70 measurements; slope 14.99 ± 44.69, p = 0.185).

These plots in Figure 4 depict the temporal dynamics of assay readings over multiple visits, revealing both cohort-level trends and substantial inter-individual variability. While mean trajectories suggest gradual increases in functional activity over time, the wide confidence intervals and divergent individual paths underscore the biologica heterogeneity inherent in ALS progression.

**Figure 4.**
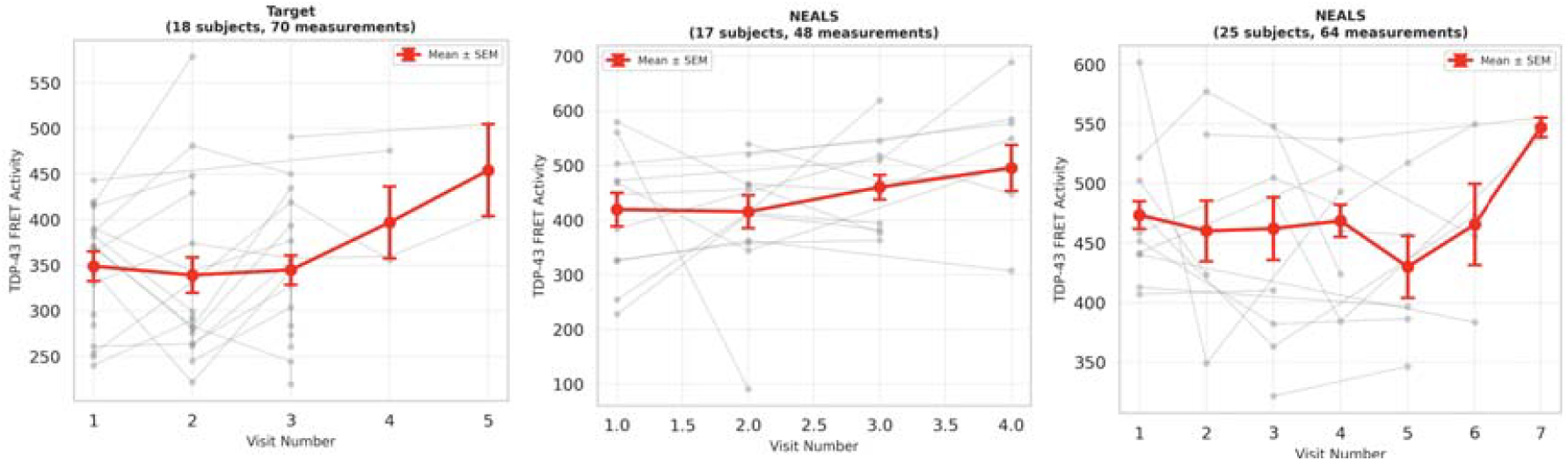
Longitudinal trajectories of TDP 43 functional activity: Temporal dynamics for Target ALS, NEALS Bio-02, NEALS Bio-03 cohorts.

Although none of these trends achieved statistical significance. As illustrated in Figure 5, the largest cohorts (Target ALS, Answer ALS, NEALS Bio-02) show positive mean slopes consistent with modest increases in TDP-43 functional activity over time, but none reached statistical significance because of large inter-individual variability within each cohort. This consistent lack of a significant longitudinal trend suggests TDP-43 signal is elevated at baseline and does not show a consistent trajectory across cohorts. For example, the time from disease onset to death, of 79 patients from the Answer ALS cohort whose time of death was reported, range from 1.3 to 14.7 years, demonstrating a longer than expected survival rate for individuals living with ALS.

**Figure 5:**
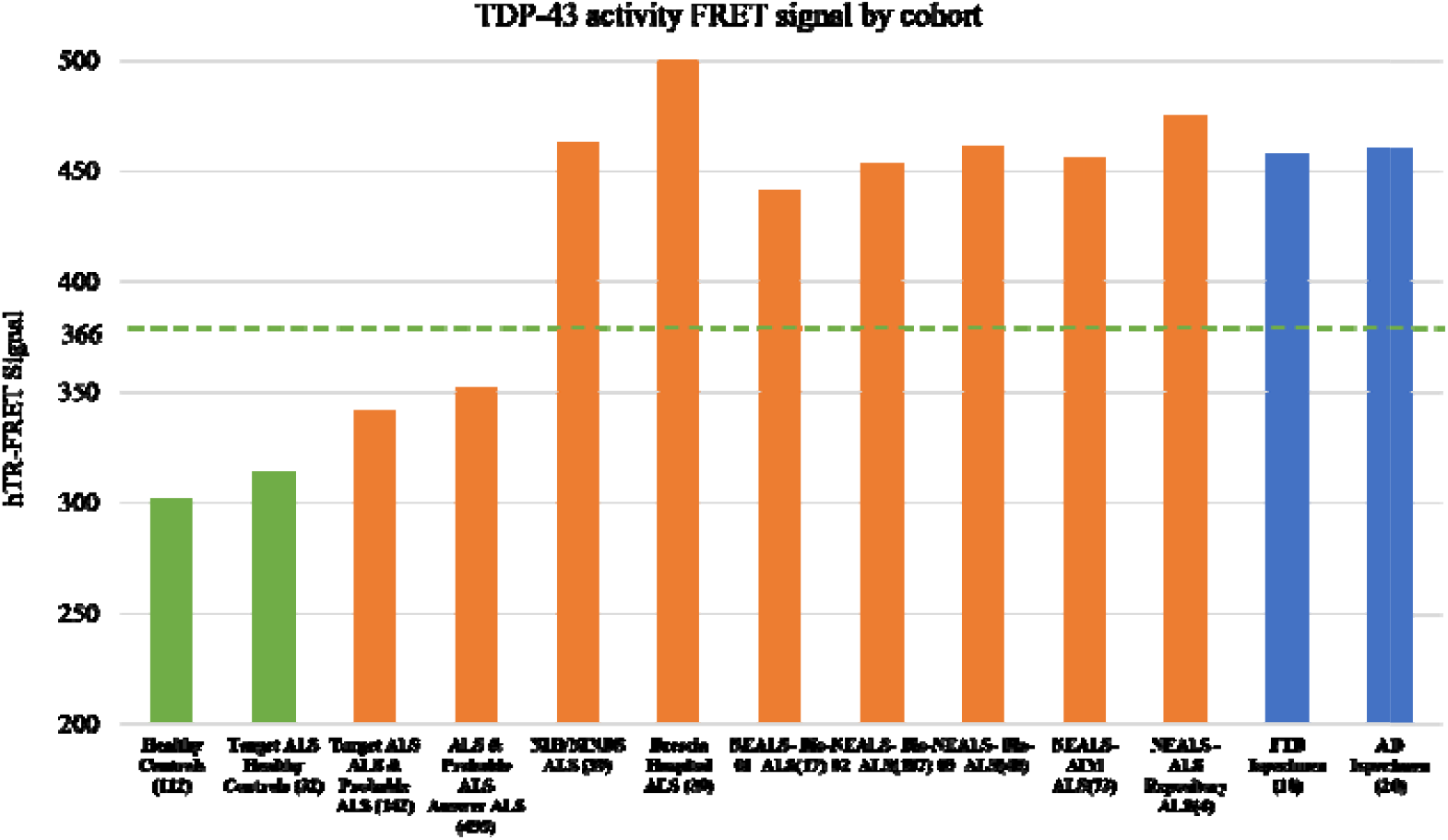
Cohort level stratification of TDP 43 functional activity: Average TDP-43 functional activity FRET values for cohort and standard deviations. Bar graph showing hTR-FRET distributions stratified by Healthy Pooled Serum (HPS), healthy Controls, ALS, FTD and AD in different colors and by contributing cohort of ALS samples (NEALS Bio01, NEALS Bio02, NEALS Bio03, NEALS AIM, NEALS ALS Repository, ANSWER ALS, Target ALS, NIH/NINDS, Spedali Civili Hosp tal of Brescia). Bar graphs display kernel density; internal box indicates median and IQR; sample counts per subgroup annotated below each bar. Healthy Pooled Serum (HPS) was used strictly as an internal plate control for normalization, not as a biological cohort. AD-fast and AD-slow were defined based on clinical progression rates and cognitive decline trajectories reported in the source biorepository.

### 3.7 Lack of Consistent Correlation with Clinical Severity (ALSFRS-R)

We next correlated baseline TDP-43 function activity with clinical severity using the ALSFRS-R score (Fig. 6). Across the cohorts, there was no consistent, significant correlation between TDP-43 function activity and ALSFRS score. For example, the NEALS - Bio-02 (n=97) and NEALS - Bio-03 (n=48) cohorts showed weak, non-significant negative trends (NEALS - Bio-02, r = -0.052, p = 0.613; NEALS - Bio-03, r = -0.013, p = 0.928). An exception was observed in the Target ALS cohort (n=63), which demonstrated a statistically significant, modest negative correlation (Pearson r = -0.3146, p = 0.012; Spea man rho = -0.2765, p = 0.0283) between ALSFRS-R and TDP-43 functional activity.

**Figure 6.**
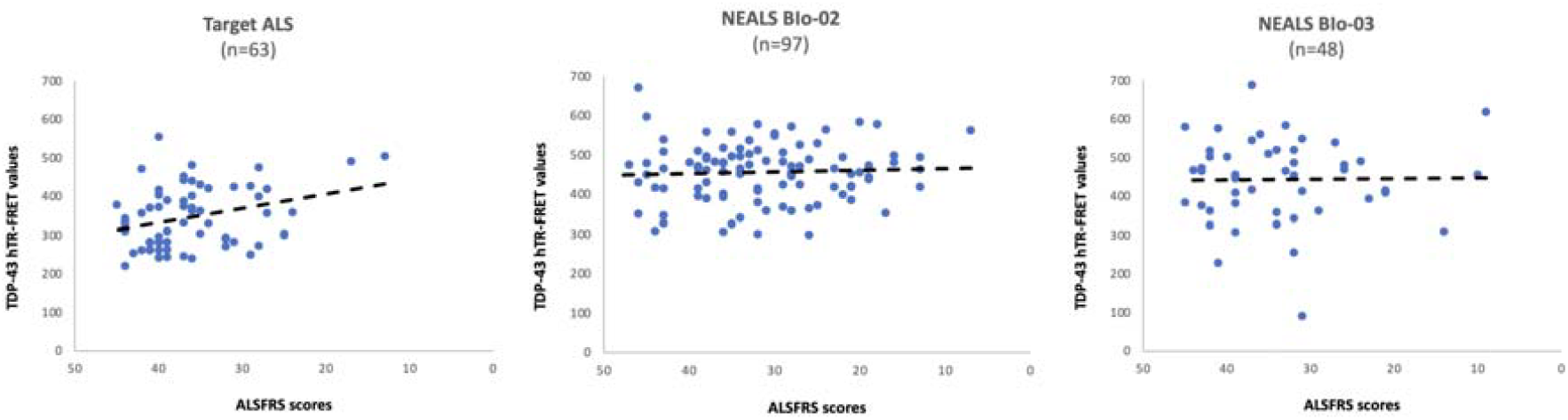
ALSFRS Correlations: Clinical severity associations for Target ALS, NEALS Bio-02, NEALS Bio-03 cohorts.

### 3.8 Weak Correlation in ANSWER ALS Highlights Cohort Heterogeneity

Similarly, when exploratory correlation value by correlating the rate of change in TDP-43 functional activity with the rate of functional decline (ALSFRS-R slope), the results were highly variable. The largest cohort, Answer ALS (n=134), showed a weak pseudo-longitudinal trend (overall R^2^ ≈ 0.07). This weak association was most pronounced in early-progression (defined as Quartile 1 based on time from symptom onset to death/last follow-up) individuals (Quartile 1, R^2^ = 0.25), (Fig. 7).

**Figure 7.**
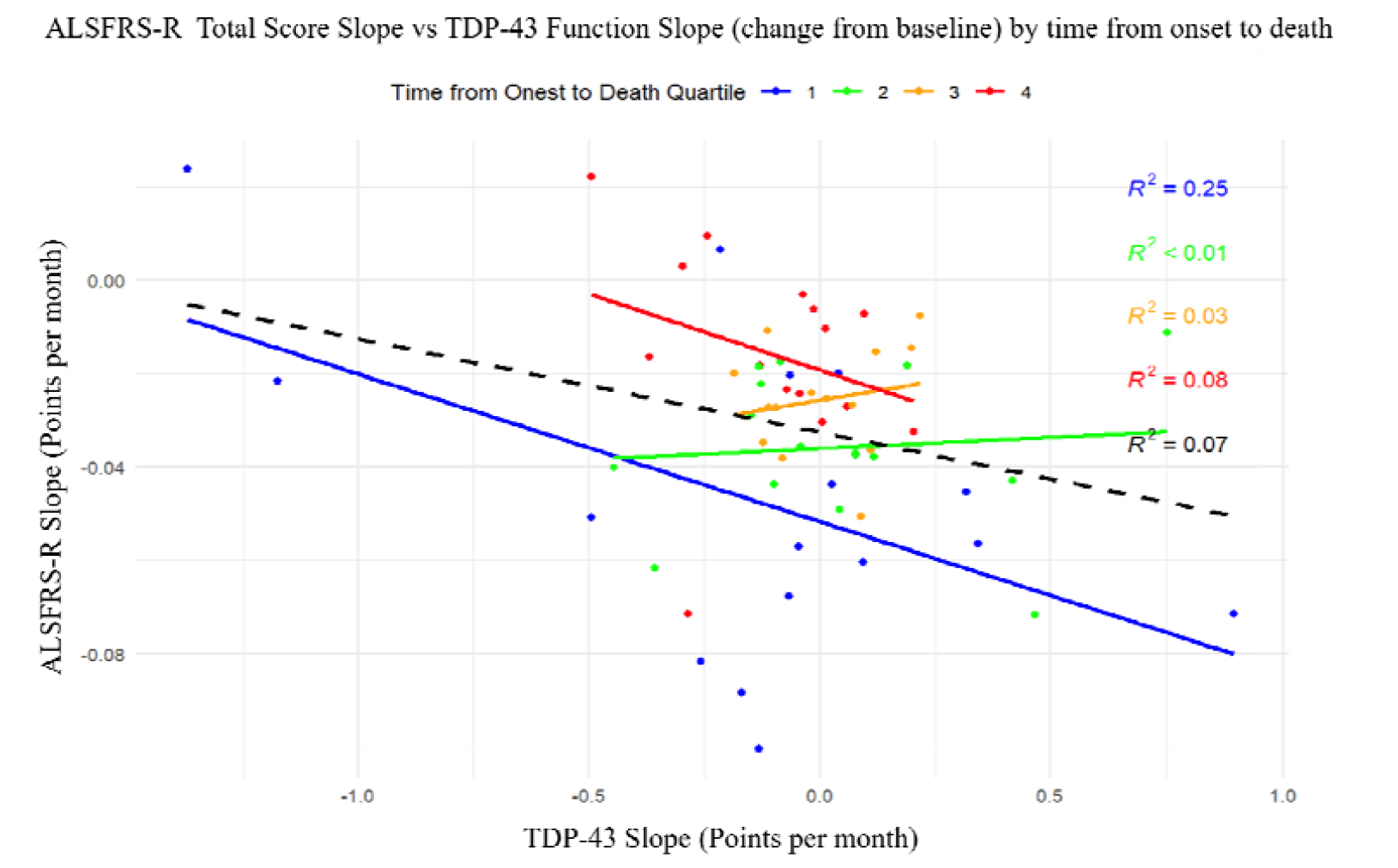
Correlation Between Functional Decline and TDP-43 hTR-FRET Values: Scatter plot of the ALSFRS-R decline rate (points/month) against the rate of change in TDP-43 functional activity (a.u./month), stratified by quartiles of time from symptom onset to death or last known follow-up. The analysis includes deceased participants with available paired longitudinal data. The overall inverse trend (black dashed line) reflects a modest association, with considerable inter-individual scatter, indicating that steeper increases in TDP-43 activity tend to coincide with faster functional decline. This association is most pronounced in the earliest disease quartile (Q1, blue; shortes time from onset), where the correlation is strongest (R^2^ = 0.25), suggesting a closer coupling between molecular dysregulation and clinical deterioration in rapid progressors.

The weak correlations likely reflect high biological heterogeneity and inclusion of atypical disease courses. This cross-sectional analysis does not constitute a prognostic study; findings are exploratory.

### 3.9 Paired Longitudinal analysis of Paired TDP-43 Functional Activity and ALSFRS-R Scores in ANSWER ALS

A total of 134 ANSWER ALS participants were assessed, of whom 105 had concurrent baseline TDP 43 functional measurements and ALSFRS R scores. A detailed demographic and longitudinal summary of this cohort is provided in Table S1. Demographics reflected typical ALS distributions (mean age 60.5 years; 60.4% male; 88% sporadic) (Table S1.). Participants were enrolled at a median of 1.7 years from symptom onset (range 0.3–15.1 years) with baseline ALSFRS R scores averaging 36 ± 7 and TDP 43 function 358.9 ± 81 a.u. The baseline ALSFRS-R score of 36 ± 7 is consistent with mild-to-moderate clinical impairment. Between baseline and the next visit, TDP 43 functional activity showed a small mean increase from 358.9□±□81.0 a.u. to 372.1□±□92.3 a.u. (+8.9□±□83.1 a.u.; n=81), whereas ALSFRS R scores declined by 6.8%□±□12.6% (n=99) over the same interval. Group-level paired trends (↑ TDP-43, ↓ ALSFRS-R) suggest concurrent biochemical and clinical changes, though individual-level correlations remain modest and are influenced by inter-assay and biological variability. At the group level, rising TDP-43 activity was observed alongside declining ALSFRS-R scores, though individual trajectories showed high variability and modest correlation (Figure 8).

**Figure 8.**
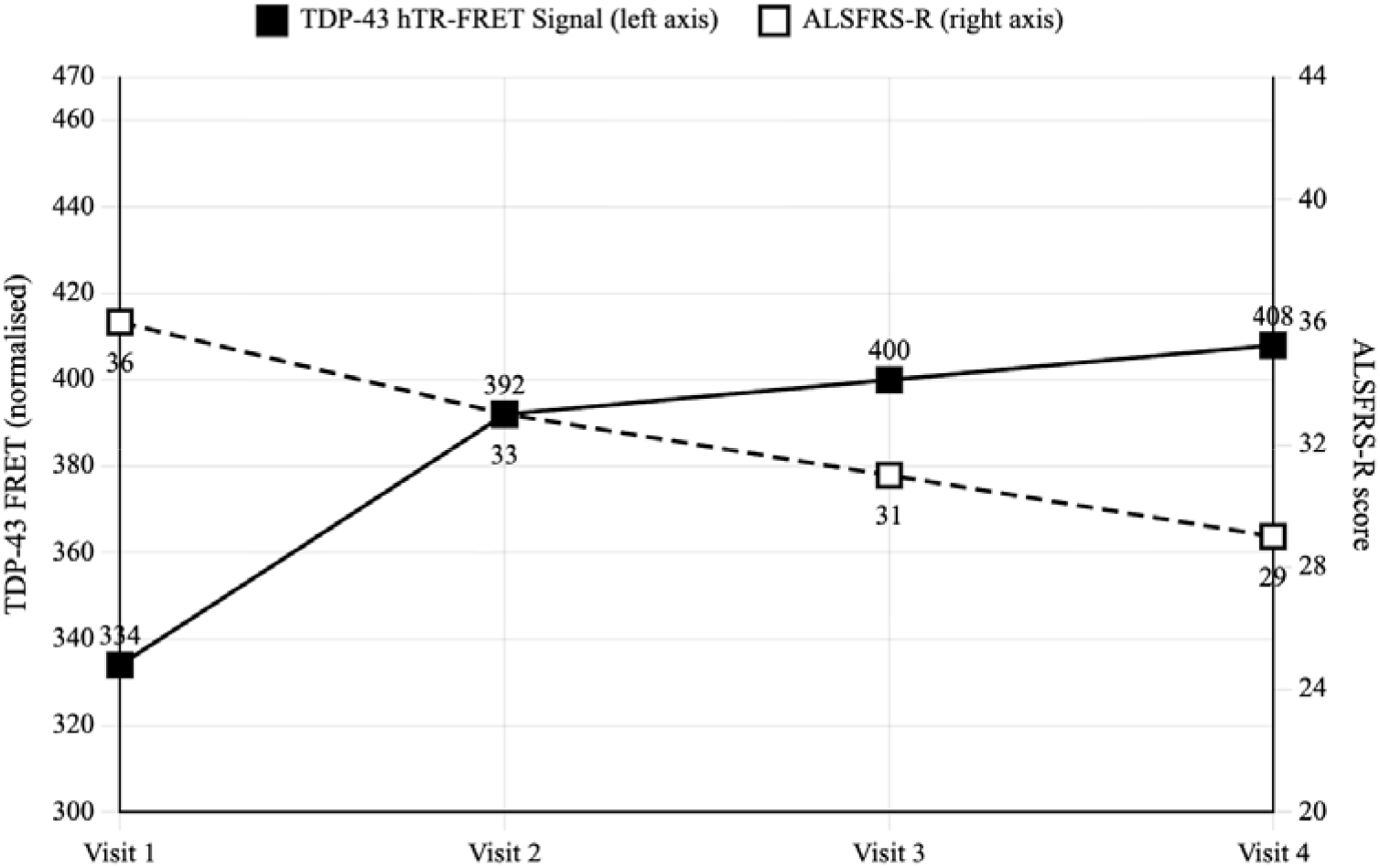
Longitudinal hTR-FRET Values and ALSFRS-R Scores Stratified by Disease Progression Quartiles: Longitudinal trajectories from ANSWER ALS participants illustrating changes in TDP-43 functional activity (a u) alongside ALSFRS-R scores over time Data are stratified by quartiles of time from symptom onset to death or last follow-up to account for disease duration heterogeneity. Group-level trends show concurrent increases in TDP-43 activity and declines in motor function, though individual trajectories exhibit substantial variability. The figure highlights the exploratory nature of these associations and the high inter-individual scatter observed in longitudinal biomarker-clinical correlations.

Genotype specific patterns were evident among 66 familial ALS samples. C9orf72 carriers exhibited a wide baseline range (≈204–600 a.u.), where higher early activity aligned with shorter survival (34–55 months), while lower or stable activity corresponded to longer survival (up to 65–68 months). SOD1 mutation carriers exhibited greater variability: participants with L114T, E100K, and c.374A>G mutations showed moderate TDP 43 functional activity with survival spanning 24–80 months. Notably, the sole SOD1 A4V carrier was a clear outlier, demonstrating low TDP-43 functional activity and preserved early ALSFRS-R despite rapid progression to death within 18 months.

## 4. Discussion

This study introduces a novel serum-based hTR-FRET assay that shifts the canonical TDP-43 biomarker development from static abundance to dynamic RNA-binding function. The functional readout discriminates ALS from controls with good classification accuracy (AUC=0.79) and shows cohort-dependent exploratory trends, where higher activity associates with worse function and shorter survival. To our knowledge, this is the first demonstration that probe-competent TDP-43 functional state can be directly quantified in human serum. Specificity is achieved by engineering the synthetic RNA binders requiring TDP-43 engagement of (UU)n RNA motifs; signal arises only when motif-competent TDP-43 bridges labeled RNA probes, effectively functioning as a molecular logic gate. Using hTR-FRET with a long-lifetime lanthanide donor (τ□ ≈ 1.6 ms) suppresses serum autofluorescence, enabling picogram sensitivity. The biphasic “dip-then-rise” trace reflects a transient initial signal decrease followed by motif-dependent rise, reporting functional TDP-43 molecules. Serum was chosen over plasma, as plasma contains anticoagulants and fibrinogen, that reduce FRET efficiency and abolish the biphasic signature [15].

### Diagnostic performance and genotype stratification

In a large cross-sectional cohort, the assay detected significantly elevated functional activity in ALS serum versus controls (AUC = 0.79). Exploratory genotype stratification showed mean values of 392 a.u. for sporadic ALS, 382 a.u. for C9orf72 ALS, and 323 a.u. for SOD1 ALS. The subgroup patterns, particularly the strong values in SOD1-ALS, suggest that TDP-43 functional activity in biofluids varies with genetic background and may reflect interactions between SOD1 pathology and TDP-43 dysfunction, however, studies need to be repeated in a larger cohort of familial ALS. Although SOD1 ALS has traditionally been considered outside the spectrum of TDP-43 proteinopathies, while emerging but not yet consensus evidence suggests that misfolded SOD1 aggregates can induce TDP-43 mislocalization and aggregation, this remains an area of active investigation and the causal directionality has not been established [16]. Additional studies have demonstrated co-deposition of SOD1 and phosphorylated TDP-43 in motor neurons of both familial and sporadic ALS cases, reinforcing the possibility of shared pathogenic cascades [17,18]. In our cohort, 28 genetically confirmed SOD1 ALS cases exhibited the hTR-FRET value (323 ± 105 a.u.), exceeding healthy controls. While exploratory due to limited sample size, these findings demonstrate that functional TDP-43 functional activity is detectable in SOD1 ALS serum and support the hypothesis that TDP-43 dysfunction may play a role even in genetically distinct ALS subtypes. The wide baseline range (≈204–600 a.u.) in C9orf72 carriers likely reflects disease heterogeneity, variable timepoints from onset, and potential differences in extracellular TDP-43 release kinetics. TDP-43 proteinopathy is a recognized hallmark in a substantial subset of FTD cases. The elevated signal likely reflects this co-pathology. In the context of emerging therapeutics targeting TDP-43 biology, a functional serum readout may prove useful for target engagement studies, pending prospective validation. The present findings are hypothesis-generating and do not yet constitute a validated pharmacodynamic tool.

### Interpreting elevated serum TDP-43 functional activity

Elevated serum hTR FRET values should not be conflated with restored nuclear splicing function. Serum TDP-43 does not behave identically to intracellular TDP-43. The assay detects probe-competent extracellular TDP-43 species released during neurodegeneration; it does not measure nuclear splicing competence or downstream transcript regulation. Thus, a higher hTR FRET value likely reflects increased probe engagement by circulating full-length or truncated TDP 43 species that preserve RBD architecture, rather than restoration of physiologic nuclear function. This distinction reconciles the apparent paradox between increased measurable activity in biofluids, and intracellular loss-of-function documented histopathologically [1,10,19].

Furthermore, several biological mechanisms may explain this increase. Neurodegeneration releases intracellular contents, including full-length and truncated TDP 43 species, into biofluids; some of these extracellular species preserve RBD architecture and transiently bind UU rich sequences [14,20]. Proteolytic cleavage and post-translational modifications produce a heterogeneous pool where a subset of molecules remains aggregation prone but probe competent. Furthermore, mislocalized cytoplasmic TDP 43 may participate in aberrant assemblies that paradoxically increase RNA-binding interface accessibility even as nuclear function collapses [21]. Therefore, a higher hTR FRET amplitude likely indexes the quantity of probe competent TDP 43 species in serum rather than an intact splicing program. Rising extracellular TDP-43 may reflect progressive cellular release and accumulation of probe-competent species as neurodegeneration advances.

### Longitudinal trends and potential as a progression biomarker

Longitudinal analysis revealed highly variable trajectories. Most longitudinal analyses were non-significant and exhibited substantial inter-individual variability. While the largest cohorts exhibited positive mean slopes, none reached statistical significance. These findings represent exploratory associations warranting prospective clinical study. C9orf72 carriers showed particularly pronounced early elevation that aligned with more aggressive disease and reduced survival, though larger cohorts are required to validate stratification utility. SOD1 cases largely followed similar trends (n=14/15 samples), but mutation-specific outliers (for example, an A4V carrier with rapid decline despite low serum activity) highlight that some pathogenic trajectories are TDP-43-independent.

This provides the first direct biofluid evidence supporting TDP-43 as a potential progression biomarker and aligns with models implicating extracellular TDP-43 in disease propagation [17,18,22]. While exploratory genotype patterns are noted, larger cohorts are required to validate stratification utility. The present findings are hypothesis-generating and do not yet constitute a validated pharmacodynamic tool. Baseline TDP-43 functional activity correlated negatively with ALSFRS-R in the Target ALS cohort, but this association was not replicated across all samples of the Answer ALS cohort, emphasizing the impact of pre-analytical variability. Factors such as sample age, freeze-thaw cycles, clotting time, and collection protocols can influence protein solubility and assay readouts [23]. We therefore interpret inter cohort value differences cautiously and recommend harmonized pre analytic SOPs and prospective stability studies to disentangle true biological signals from handling artifacts. While NEALS employs standardized SOPs, differences in site-specific handling, storage duration, and assay batch effects can still introduce variability. Harmonization remains a key recommendation for future multi-cohort integration. NEALS SOPs represent best practice but were not uniformly applied in all samples (e.g., iSpecimen-sourced AD/FTD comparator samples). A formal pre-analytic stability study is proposed as a priority for the next phase. A formal Discovery/Replication design is required to resolve cohort-specific slope differences, which may reflect assay noise and limited sample sizes rather than true biological differences. The weak correlations likely reflect high biological heterogeneity, atypical disease courses, and pre-analytic variability, which mask clearer associations. Longitudinal associations remain exploratory and highly variable, warranting prospective validation. The claim of “strong prognostic value” is not supported by the current data. SOD1 pathology may indirectly influence TDP-43 dynamics, but mutation-specific outliers (e.g., A4V) highlight TDP-43-independent progression pathways. At the group level, paired trends (↑ TDP-43, ↓ ALSFRS-R) suggest concurrent biochemical and clinical changes, though individual-level correlations remain modest. High biological heterogeneity, pre-analytic variability, and atypical disease courses likely mask clearer associations. Longitudinal associations remain exploratory and warrant prospective validation.

In summary, elevated hTR FRET value in ALS most plausibly reflects an increased burden of dysfunctional extracellular, probe competent TDP 43 species. In the context of emerging therapeutics targeting TDP-43 biology, a functional serum readout may prove useful for target engagement studies, pending prospective validation.

## 5. Conclusion

The hTR-FRET TDP-43 functional assay represents a paradigm shift from measuring the static presence of a pathological protein to quantifying its dynamic biochemical function. By integrating sophisticated RNA-probe design with highly sensitive hTR-FRET detection, we have developed a serum-based assay that demonstrates moderate group-level discrimination of ALS from controls and exploratory associations with disease trajectory, providing a proof-of-concept platform warranting prospective clinical study.

## Data Availability

All data produced in this study are available from the authors upon reasonable request. Serum samples were obtained from NEALS, NIH/NINDS, Answer ALS, Target ALS, and other biorepositories under their respective data-use agreements. Processed hTR‑FRET values and analysis code can be shared upon request.

## Author Contributions

K.S.S., V.L.D., and I.T. led the conceptualization, data analysis, and manuscript drafting. J.C., H.S., and E.G. performed the experiments and generated the primary data. I.T. and T.H-B. designed and validated the RNA substrates. E.B., G.S.V., M.E.C., J.B., B.F., A.N., J.R., R.B., B.B., and A.P. coordinated clinical cohort recruitment and sample management. A.N. and R.B. oversaw biorepository operations and data integration. All authors reviewed and approved the final manuscript.

## Institutional Review Board Statement

This study received ethical approval from the Mass General Brigham IRB (protocol 2006P000982), the Barrow Neurological Institute DHARE IRB (approval PHOENIX 21 500 101 70 09), the NIH/NINDS IRB, and the Ethics Committee of ASST Spedali Civili di Brescia, Italy (approval NP 5285). TARGET ALS and the local IRBs for each contributing cohort also provided approval. All procedures involving human participants were conducted in accordance with institutional and national ethical standards and the Declaration of Helsinki and its later amendments.

## Informed Consent Statement

All participants (or their legal guardians, where applicable) underwent an informed consent process and provided written informed consent prior to sample collection and data use.

## Acknowledgements

We thank the NEALS Consortium for its leadership in biobanking and for recognizing initial findings of this research with the 2024 Basic Science Award. We are deeply grateful to the patients and families whose biospecimens made this research possible. Additional support was provided by the NIH/NINDS Intramural Research Program (ZIA NS 003130) and through collaborations with Massachusetts General Hospital, Barrow Neurological Institute, the International Centre for Genetic Engineering and Biotechnology (ICGEB), Trieste, and the Spedali Civili Hospital of Brescia. We sincerely thank Hillel Bachrach (Nemdx) for scientific discussions that strengthened the conceptual framework; Dalia Cohen and Zafrira Avnur (Nemdx) for critical manuscript review, Hong Li for developing an epidemiological study design; Alexandra Lea Kipping, Luis Holdsworth for their dedicated technical support and Rebecca Bornheimer for bioanalytical analysis of the ANSWER-ALS cohort. We also acknowledge Amy Easton and Laura Dugom (Target ALS) for their support and constructive feedback. The contributions of the NIH author(s) were made as part of their official duties as NIH federal employees, are in compliance with agency policy requirements, and are considered Works of the United States Government. However, the findings and conclusions presented in this paper are those of the author(s) and do not necessarily reflect the views of the NIH or the U.S. Department of Health and Human Services.

## Data Availability Statement

Details of the probe sequences and assay design are disclosed in the patent applications WO2024115914A1 and EP4347873A1. Source data for the supplementary materials are included online. Supplementary Data 1 contains the data underlying Table S1, and Supplementary Data 2 contains the data underlying Figure S1.

